# Is an earlier onset of focal epilepsy associated with atypical language lateralization? A systematic review, meta-analysis and new data

**DOI:** 10.1101/2024.11.13.24315462

**Authors:** Freya Prentice, Lara Chehabeddine, Maria Helena Eriksson, Jennifer Murphy, Leigh N. Sepeta, William D. Gaillard, Madison M. Berl, Frédérique Liégeois, Torsten Baldeweg

## Abstract

Right and bilateral language representation is common in focal epilepsy, possibly reflecting the influence of epileptogenic lesions and/or seizure activity in the left hemisphere. Atypical language lateralization is assumed to be more likely in cases of early seizure onset, due to greater language plasticity in childhood. However, evidence for this association is mixed, with most research based on small samples and heterogenous cohorts. In this preregistered meta-analysis we examined the association between age at seizure onset and fMRI-derived language lateralization in individuals with focal epilepsy. The pooled effect size demonstrated a correlation between an earlier onset and rightward language lateralization in the total sample (r=0.1, p=.005, k=58, n=1240), with no difference in the correlation between left and right hemisphere epilepsy samples (Q=62.03, p=.302). In exploratory analyses of the individual participant data (n=1157), we demonstrated strong evidence that a logarithmic model fits the data better than a linear (BF=350) or categorical model with 6 years of age as a cut-off (BF=36). These findings indicate that there is a small but significant relationship between age at seizure onset and language lateralization. The relationship was consistent with theories of language plasticity proposing an exponential decline in plasticity over early childhood. However, given that this effect was subtle and only found in larger sample sizes, an early age at seizure onset would not serve as a good indicator of atypical language lateralization on the individual patient level.

## 1. Introduction

Most of the general adult population is left lateralized for language (approximately 95% of right-handers: Knecht et al., 2000; Springer et al., 1999; and 75% of left-handers: Szaflarski et al., 2002), and left language lateralization is present from early childhood (Olulade et al., 2020). Children and adults with focal epilepsy, however, show higher rates of right and bilateral patterns of language representation compared to those seen in the general population (Berl, Zimmaro, et al., 2014; Rasmussen & Milner, 1977). This is presumed to reflect disruption of the typical trajectory of left language specialization or potential interhemispheric reorganisation of language networks from the left to the right hemisphere (as first suggested in early case series; Basser, 1962; Lenneberg, 1967; Rasmussen & Milner, 1977). Atypical language lateralization has been associated with disruption from epileptogenic lesions (Liégeois et al., 2004; Weber et al., 2006) and seizure activity (Berl et al., 2005; Branch et al., 1964; Janszky et al., 2006) in the left hemisphere.

Atypical language lateralization in focal epilepsy has also been associated with an earlier age at seizure onset in studies using both intracarotid amobarbital procedure (Brazdil et al., 2005) and functional magnetic resonance imaging (fMRI) (Anderson et al., 2006; Gaillard et al., 2007; Springer et al., 1999; Woermann et al., 2003). This association may be specific to those with epilepsy in the left hemisphere (Helmstaedter et al., 1997; Rey et al., 1988). Such findings fit with theories of language plasticity, which describe trajectories of increasing specialization and lateralization of language networks over childhood, accompanied by decreasing plasticity (Lenneberg, 1967; Olulade et al., 2020; Satz et al., 1990; Vargha-Khadem et al., 2000). Accordingly, reorganisation of language from the left to right hemisphere would be more likely when an insult is sustained at an early age and the likelihood would decrease over development Based on early case series, Lenneberg (1967) suggested that there was a critical period for language reorganisation between the ages of 2 and 14 years, and that injury after this period would be unlikely to trigger reorganisation. A critical or sensitive period for language reorganisation has remained a popular idea, with more recent studies in children with intractable epilepsy indicating that 6 years of age may be the potential ‘boundary’ before which plasticity is greater, and after which reorganisation is less likely (Saltzman-Benaiah et al., 2003). Complementary evidence comes from the stroke literature where studies have found that atypical language lateralization is associated with an earlier age at insult to the left hemisphere (Szaflarski et al., 2014). Right and bilateral language representation appear most common when the stroke occurs before the age of 2 years, or between 2 and 5 years, respectively (Lidzba et al., 2017). In contrast, others have suggested that instead of a sharp cut-off there is a gradual linear decline in the proportion of individuals with reorganisation as age at seizure onset increases (Helmstaedter et al., 1997; Springer et al., 1999), and therefore the relationship between age at seizure onset and language lateralization is linear.

Some studies have failed to identify a significant association between age at seizure onset and language lateralization (Janszky et al., 2003; Janszky et al., 2006; Sveller et al., 2006; You et al., 2011). Inconsistent findings may partially be driven by methodological differences between studies, specifically the different language tasks and regions of interest (ROI) used for the calculation of laterality indices (LI) from fMRI. Crossed lateralization, where language functions supported by frontal and temporal regions are supported by the opposite hemispheres, is more common in epilepsy than in the general population (Berl, Zimmaro, et al., 2014). In addition, Duke et al. (2012) found that temporal lobe epilepsy was associated with atypical language lateralization in temporal but not frontal ROIs, perhaps suggesting that the former may be more predisposed to reorganisation. Therefore, whether age at seizure onset is associated with language lateralization may depend on whether lateralization is measured in frontal or temporal ROI.

Given the heterogeneity of the current literature, a meta-analysis is needed to address whether there is an association between age at seizure onset and language lateralization in focal epilepsy, and whether this relationship is linear or suggests a ‘sensitive period’ hypothesis. To disentangle the heterogenous populations and methodologies, we examined how this association varied based on the side (left vs. right) and location (frontal vs. temporal) of epilepsy pathology, and ROI chosen (frontal vs. temporal) for LI calculation.

## 2. Method

### 2.1 Preregistration

We conducted a systematic literature search following the 2020 Preferred Reporting Items for Systematic Review and Meta-Analysis (PRISMA) guidelines (Page et al., 2021). The preregistration protocol was uploaded to the Open Science Framework on 19^th^ May 2022 before data extraction began and can be found at this link: https://tinyurl.com/bdctb3t2. The preregistration outlines the search strategy, study selection, data extraction and data synthesis processes. The completed PRISMA checklists, summary statistics for each identified study and meta-analysis script can also be found at this link.

### 2.2 Search and screening

We searched Embase, MEDLINE and PsycInfo databases for studies that were available online before August 14^th^ 2024^1^ using the keywords, MeSH and Embase terms in Table 1. Keywords, MeSH and Embase terms within a concept were combined with ‘OR’ and the different concepts were combined using ‘AND’. The search terms were the same as outlined in the preregistration but with the addition of MeSH and Embase terms and the term ‘Magnetic Resonance Imaging’. We carried out an additional citation search by screening all articles which cited influential studies on the topics of LI methodology (Adcock et al., 2003; Baciu et al., 2005; Holland et al., 2001; Jansen et al., 2006; Liégeois et al., 2002; Nagata et al., 2001; Seghier, 2008; Wilke & Lidzba, 2007) and early fMRI language lateralization (Hertz-Pannier et al., 1997; Springer et al., 1999).

**Table 1.** Keywords, MeSh and Embase terms used in literature search

| Concept | Keywords | MeSH or Embase terms |
| --- | --- | --- |
| Epilepsy | epilep* OR epilepsy | Medline: Epilepsy<br>Embase: Epilepsy |
| Language | language OR linguistic* OR verbal<br>OR speech | Medline: Language<br>Embase: Language |
| fMRI | magnetic resonance imaging OR<br>functional magnetic resonance<br>imaging OR functional MRI OR fMRI | Medline: Magnetic resonance imaging<br>Embase: Functional magnetic resonance<br>imaging |

Two independent reviewers (F.P. and L.C.) examined the identified titles, abstracts and studies based on the exclusion criteria (see Table 2). Disagreement when screening was resolved through discussion with a third reviewer (T.B.). A total of 34 articles were included in the final meta-analysis. A flow chart showing how many studies were identified and excluded at each stage is shown in Fig. 1.

**Table 2.**
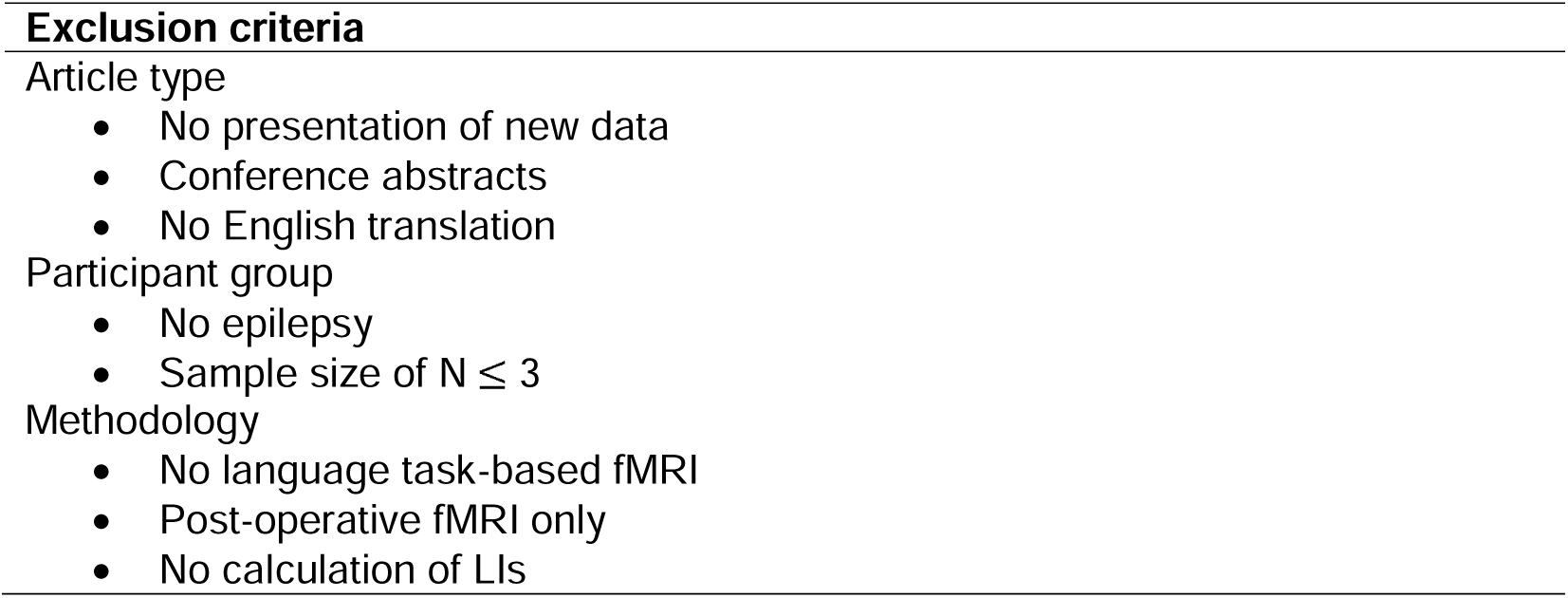
Exclusion criteria for meta-analysis

| **Exclusion criteria** |
| --- |
| Article type |
| • No presentation of new data<br>• Conference abstracts<br>• No English translation |
| Participant group |
| • No epilepsy<br>• Sample size of $N \leq 3$ |
| Methodology |
| • No language task-based fMRI<br>• Post-operative fMRI only<br>• No calculation of LIs |

**Fig. 1.**
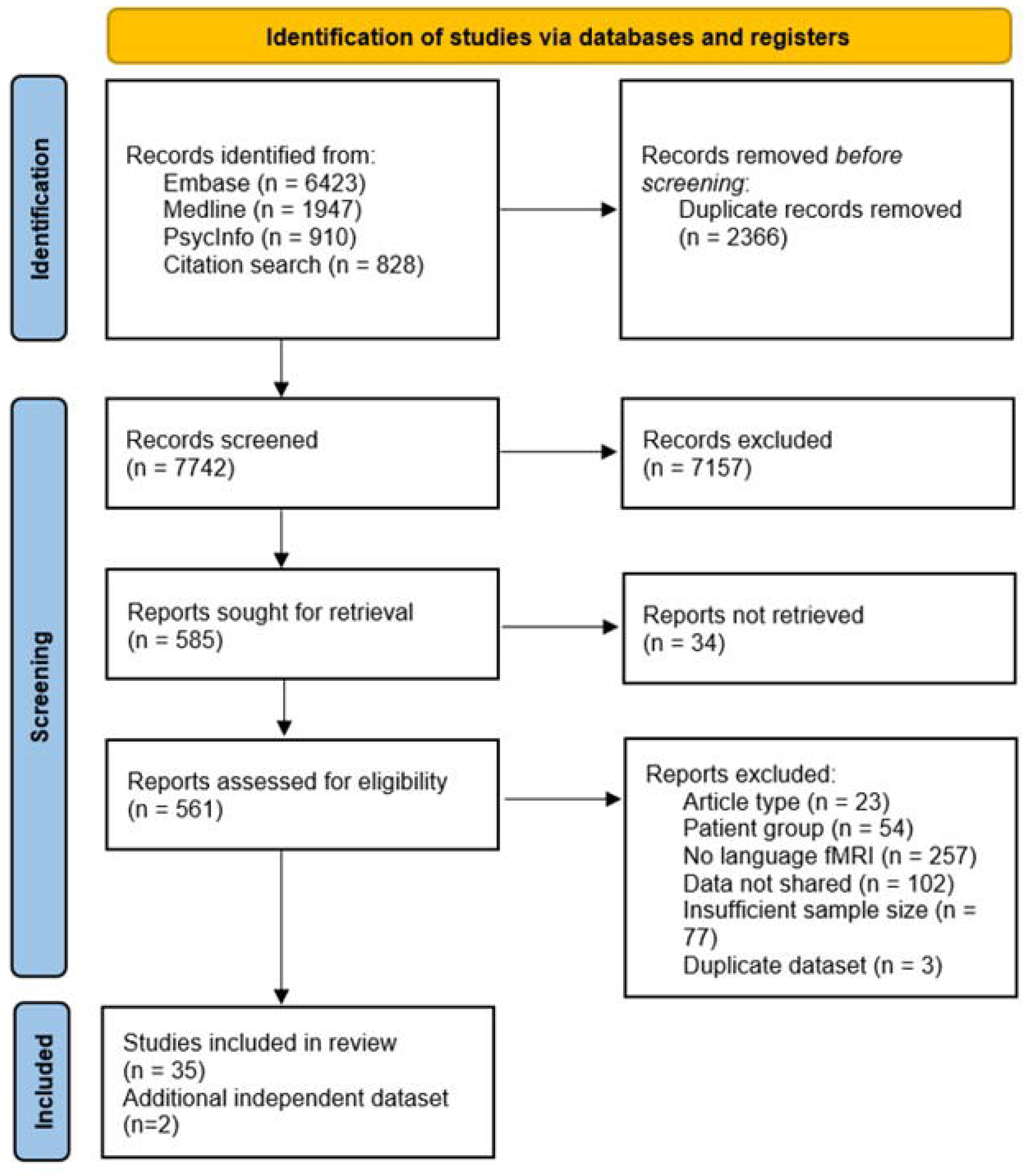
Flow chart depicting the number of published reports identified by the database and citation searches and how many were excluded at each stage of the screening process.

### 2.3 Exclusion criteria

The exclusion criteria can be found in Table 2. The exclusion differed from the preregistration in two ways. First, studies were excluded if the sample size was less than or equal to three, rather than less than or equal to two, as more than three participants are required to estimate the sampling variance of the effect size for each study. Secondly, the exclusion criteria of individuals being under 18 years of age at the time of seizure onset was removed. The later exclusion criterion was originally proposed because a linear relationship was not expected between age at onset and LIs in adulthood when plasticity would be greatly reduced compared to in (early) childhood. As this exclusion criterion substantially reduced the sample size and resulted in the exclusion of several articles, it was removed, and the meta-analysis was run on individuals with an age at onset in adulthood as well as childhood. We reran the meta-analyses with the original exclusion criterion of onset <18 years of age and the results were broadly consistent with the results without this exclusion criterion. This can be found in the Supplementary Materials.

### 2.4 Inclusion of additional samples

Retrospective data from two centers were added to the data collected as part of the meta-analysis. The addition of these two cohorts increased the sample size to 1254 participants (k=37). 14 of these participants were not included in the meta-analysis as they represented a sample size of 3 or less participants for the right-sided sample (Genetti et al., 2013; Herfurth et al., 2022; Hertz-Pannier et al., 1997; Koop et al., 2021; van der Kallen et al., 1998; Wilke et al., 2011). Consequently, the total sample size for meta-analysis was 1240.

#### 2.4.1 Great Ormond Street Hospital

We included a retrospective cohort of 268 children who underwent language task-based fMRI as part of their presurgical evaluation at Great Ormond Street Hospital (GOSH) in London, UK. We screened all children who had language fMRI at GOSH from 2000 to 2022 (N=350). Participants were excluded if fMRI was unavailable or poor quality or LI calculation failed (30), age at seizure onset was missing (16) or epilepsy was not lateralized (36). Due to the inclusion of this dataset, we excluded data from a previously published study that contained a partially overlapping sample (Pahs et al., 2013). Ethical approval was granted by the Great Ormond Street Hospital clinical audit department as a service evaluation (No. 1443, extended), according to the guidelines set by NHS Research Ethics Committee Review.

#### 2.4.2 Children’s National Hospital

We also included a retrospective cohort of 175 children who underwent language task-based fMRI as part of their presurgical evaluation or a research study at the Children’s National Hospital (CNH) in Washington, DC, US, from 2003 to 2023. We screened 305 children who had fMRI at CNH. Participants were excluded if fMRI was unavailable or poor quality or LI calculation failed (75), age at seizure onset was missing (5) or epilepsy was not lateralized (50). Participants were part of clinical or research protocols with different aims related to pediatric epilepsy and language development, but all were approved by the Institutional Review Board at CNH. Parents provided informed written consent and all children gave assent to participate.

### 2.5 Data extraction

The data extracted for each included study are summarized in Table 3. Data were extracted from included studies by F.P. and checked by L.C. Where data were not available in the main manuscript or supplementary material, the data were requested from the corresponding author. Of the included articles, two reported correlation coefficients for the correlation between age at seizure onset and language lateralization, 27 reported individual participant data in the article or supplement, and for five article the individual participant data were shared by the authors. Table 3 outlines how data were extracted for each included study. Where individual participant data were available, participants were excluded from the total sample if (1) the side of epilepsy, age at seizure onset or LIs were unavailable, or (2) if they were part of another included sample. A list of how many individuals were removed from each study and the reason for exclusion is included in Supplementary Table 1.

**Table 3.**
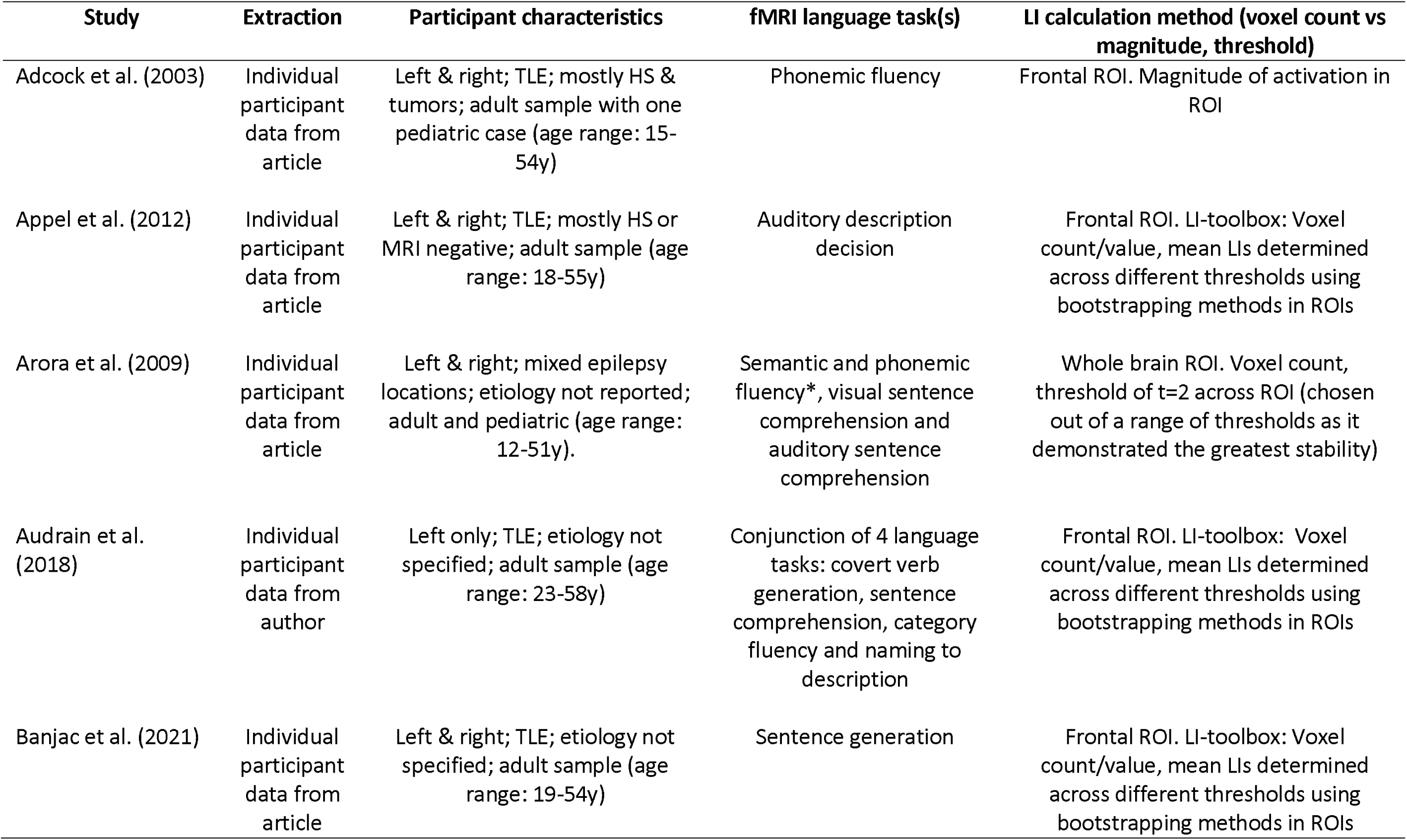
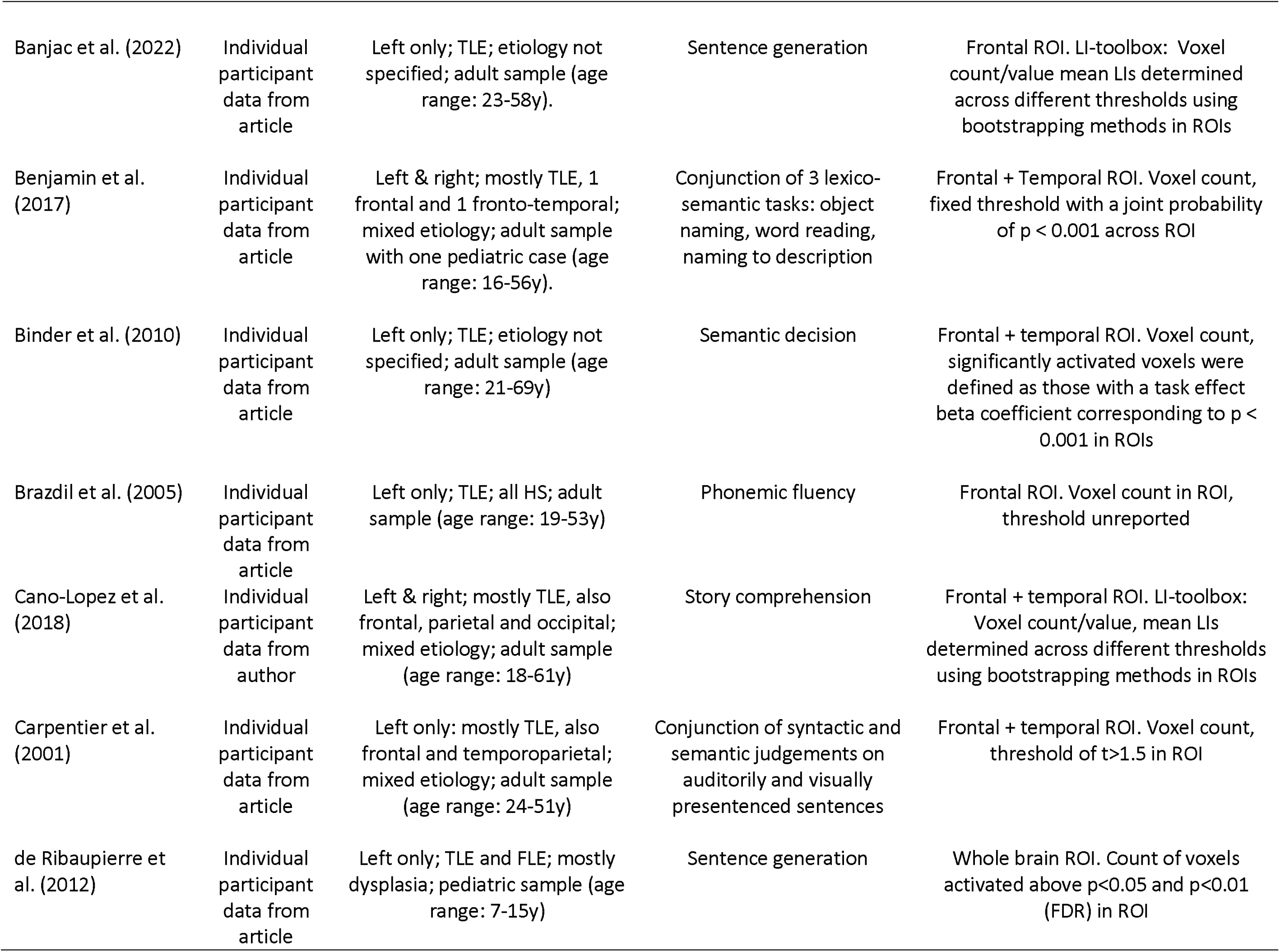
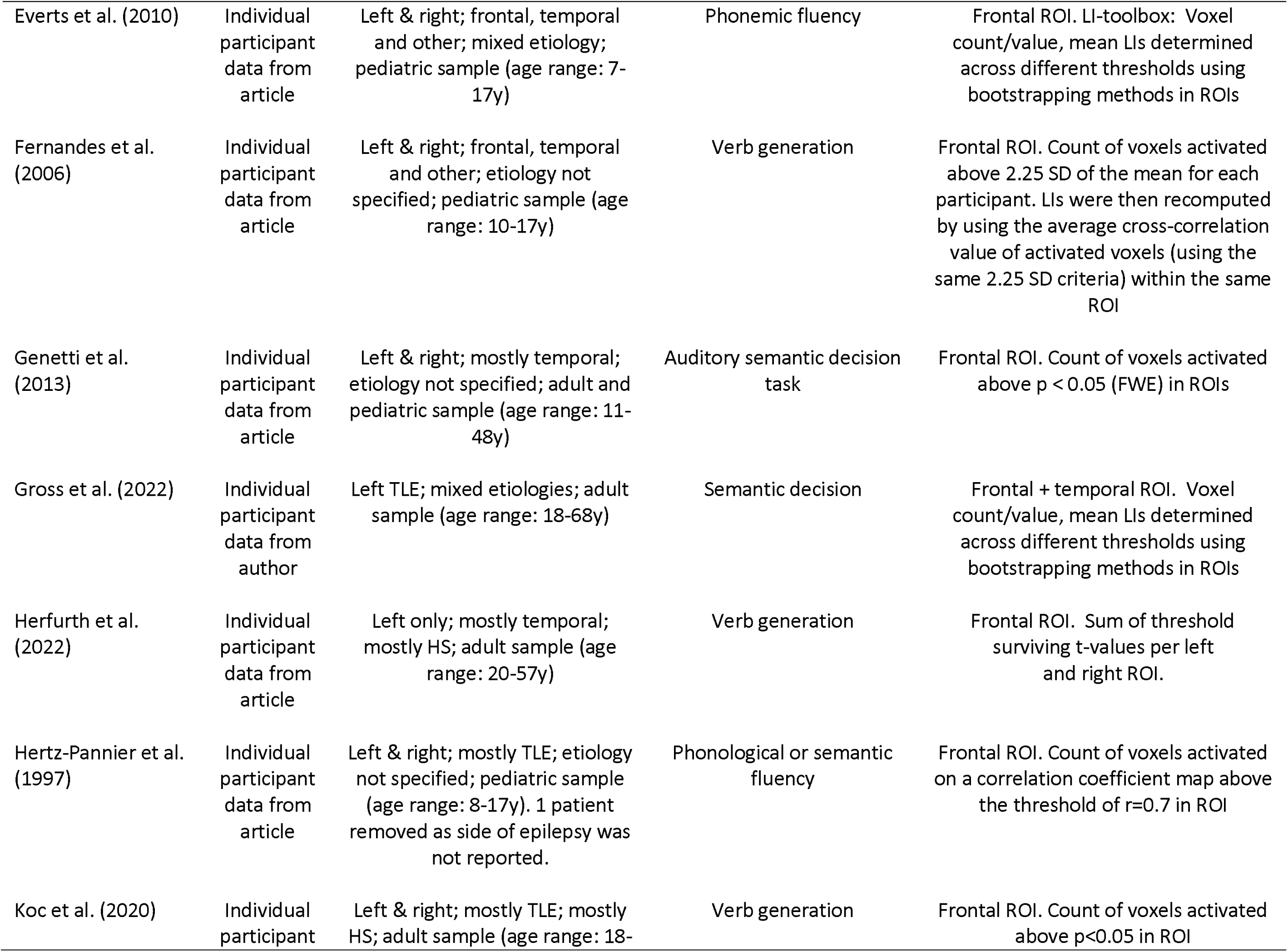
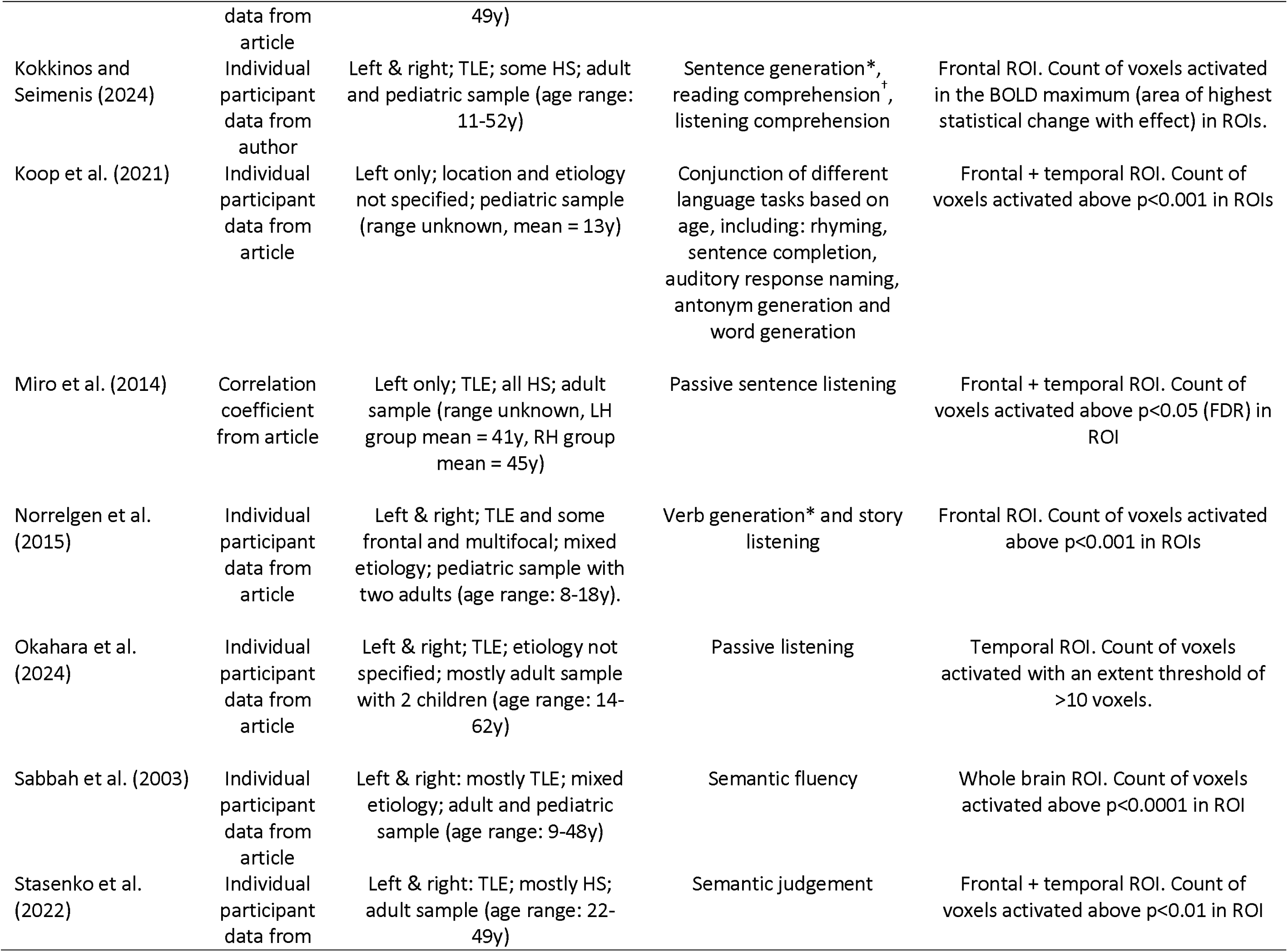
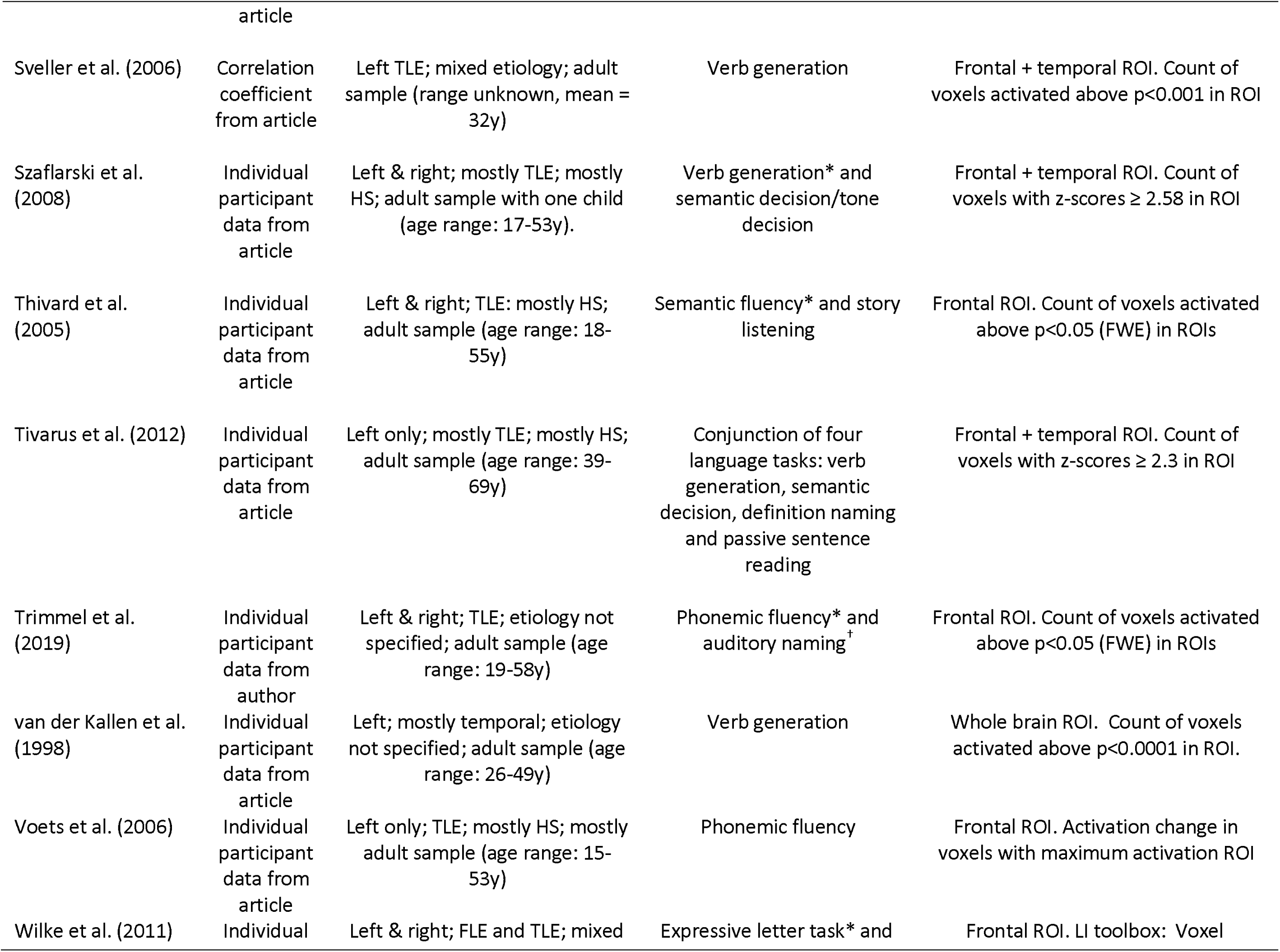
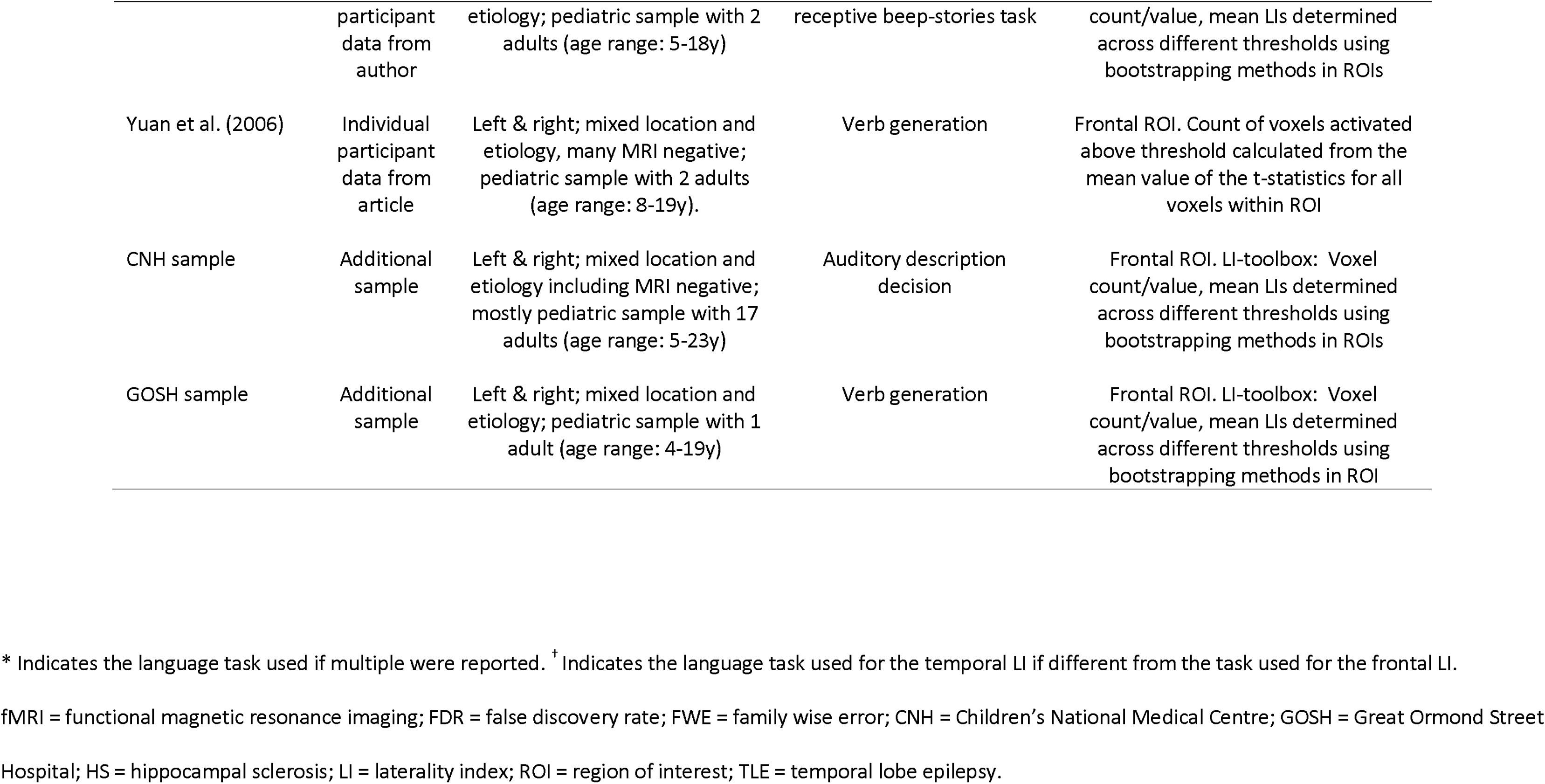
Characteristics of studies included in meta-analysis

| Study | Extraction | Participant characteristics | fMRI language task(s) | LI calculation method (voxel count vs magnitude, threshold) |
| --- | --- | --- | --- | --- |
| Adcock et al. (2003) | Individual participant data from article | Left & right; TLE; mostly HS & tumors; adult sample with one pediatric case (age range: 15-54y) | Phonemic fluency | Frontal ROI. Magnitude of activation in ROI |
| Appel et al. (2012) | Individual participant data from article | Left & right; TLE; mostly HS or MRI negative; adult sample (age range: 18-55y) | Auditory description decision | Frontal ROI. LI-toolbox: Voxel count/value, mean LIs determined across different thresholds using bootstrapping methods in ROIs |
| Arora et al. (2009) | Individual participant data from article | Left & right; mixed epilepsy locations; etiology not reported; adult and pediatric (age range: 12-51y). | Semantic and phonemic fluency*, visual sentence comprehension and auditory sentence comprehension | Whole brain ROI. Voxel count, threshold of $t=2$ across ROI (chosen out of a range of thresholds as it demonstrated the greatest stability) |
| Audrain et al. (2018) | Individual participant data from author | Left only; TLE; etiology not specified; adult sample (age range: 23-58y) | Conjunction of 4 language tasks: covert verb generation, sentence comprehension, category fluency and naming to description | Frontal ROI. LI-toolbox: Voxel count/value, mean LIs determined across different thresholds using bootstrapping methods in ROIs |
| Banjac et al. (2021) | Individual participant data from article | Left & right; TLE; etiology not specified; adult sample (age range: 19-54y) | Sentence generation | Frontal ROI. LI-toolbox: Voxel count/value, mean LIs determined across different thresholds using bootstrapping methods in ROIs |
| Banjac et al. (2022) | Individual participant data from article | Left only; TLE; etiology not specified; adult sample (age range: 23-58y). | Sentence generation | Frontal ROI. LI-toolbox: Voxel count/value mean LIs determined across different thresholds using bootstrapping methods in ROIs |
| Benjamin et al. (2017) | Individual participant data from article | Left & right; mostly TLE, 1 frontal and 1 fronto-temporal; mixed etiology; adult sample with one pediatric case (age range: 16-56y). | Conjunction of 3 lexico-semantic tasks: object naming, word reading, naming to description | Frontal + Temporal ROI. Voxel count, fixed threshold with a joint probability of $p < 0.001$ across ROI |
| Binder et al. (2010) | Individual participant data from article | Left only; TLE; etiology not specified; adult sample (age range: 21-69y) | Semantic decision | Frontal + temporal ROI. Voxel count, significantly activated voxels were defined as those with a task effect beta coefficient corresponding to $p < 0.001$ in ROIs |
| Brazdil et al. (2005) | Individual participant data from article | Left only; TLE; all HS; adult sample (age range: 19-53y) | Phonemic fluency | Frontal ROI. Voxel count in ROI, threshold unreported |
| Cano-Lopez et al. (2018) | Individual participant data from author | Left & right; mostly TLE, also frontal, parietal and occipital; mixed etiology; adult sample (age range: 18-61y) | Story comprehension | Frontal + temporal ROI. LI-toolbox: Voxel count/value, mean LIs determined across different thresholds using bootstrapping methods in ROIs |
| Carpentier et al. (2001) | Individual participant data from article | Left only: mostly TLE, also frontal and temporoparietal; mixed etiology; adult sample (age range: 24-51y) | Conjunction of syntactic and semantic judgements on auditorily and visually presented sentences | Frontal + temporal ROI. Voxel count, threshold of $t > 1.5$ in ROI |
| de Ribaupierre et al. (2012) | Individual participant data from article | Left only; TLE and FLE; mostly dysplasia; pediatric sample (age range: 7-15y) | Sentence generation | Whole brain ROI. Count of voxels activated above $p < 0.05$ and $p < 0.01$ (FDR) in ROI |
| Everts et al. (2010) | Individual participant data from article | Left & right; frontal, temporal and other; mixed etiology; pediatric sample (age range: 7-17y) | Phonemic fluency | Frontal ROI. LI-toolbox: Voxel count/value, mean LIs determined across different thresholds using bootstrapping methods in ROIs |
| Fernandes et al. (2006) | Individual participant data from article | Left & right; frontal, temporal and other; etiology not specified; pediatric sample (age range: 10-17y) | Verb generation | Frontal ROI. Count of voxels activated above 2.25 SD of the mean for each participant. LIs were then recomputed by using the average cross-correlation value of activated voxels (using the same 2.25 SD criteria) within the same ROI |
| Genetti et al. (2013) | Individual participant data from article | Left & right; mostly temporal; etiology not specified; adult and pediatric sample (age range: 11-48y) | Auditory semantic decision task | Frontal ROI. Count of voxels activated above $p < 0.05$ (FWE) in ROIs |
| Gross et al. (2022) | Individual participant data from author | Left TLE; mixed etiologies; adult sample (age range: 18-68y) | Semantic decision | Frontal + temporal ROI. Voxel count/value, mean LIs determined across different thresholds using bootstrapping methods in ROIs |
| Herfurth et al. (2022) | Individual participant data from article | Left only; mostly temporal; mostly HS; adult sample (age range: 20-57y) | Verb generation | Frontal ROI. Sum of threshold surviving t-values per left and right ROI. |
| Hertz-Pannier et al. (1997) | Individual participant data from article | Left & right; mostly TLE; etiology not specified; pediatric sample (age range: 8-17y). 1 patient removed as side of epilepsy was not reported. | Phonological or semantic fluency | Frontal ROI. Count of voxels activated on a correlation coefficient map above the threshold of $r=0.7$ in ROI |
| Koc et al. (2020) | Individual participant | Left & right; mostly TLE; mostly HS; adult sample (age range: 18- | Verb generation | Frontal ROI. Count of voxels activated above $p<0.05$ in ROI |

AGE AT SEIZURE ONSET AND LANGUAGE LATERALIZATION
|  | data from | 49y) |  |  |
| --- | --- | --- | --- | --- |
| Kokkinos and Seimenis (2024) | Individual participant data from article | Left & right; TLE; some HS; adult and pediatric sample (age range: 11-52y) | Sentence generation*, reading comprehension<sup>†</sup>, listening comprehension | Frontal ROI. Count of voxels activated in the BOLD maximum (area of highest statistical change with effect) in ROIs. |
| Koop et al. (2021) | Individual participant data from article | Left only; location and etiology not specified; pediatric sample (range unknown, mean = 13y) | Conjunction of different language tasks based on age, including: rhyming, sentence completion, auditory response naming, antonym generation and word generation | Frontal + temporal ROI. Count of voxels activated above $p < 0.001$ in ROIs |
| Miro et al. (2014) | Correlation coefficient from article | Left only; TLE; all HS; adult sample (range unknown, LH group mean = 41y, RH group mean = 45y) | Passive sentence listening | Frontal + temporal ROI. Count of voxels activated above $p < 0.05$ (FDR) in ROI |
| Norrelgen et al. (2015) | Individual participant data from article | Left & right; TLE and some frontal and multifocal; mixed etiology; pediatric sample with two adults (age range: 8-18y). | Verb generation* and story listening | Frontal ROI. Count of voxels activated above $p < 0.001$ in ROIs |
| Okahara et al. (2024) | Individual participant data from article | Left & right; TLE; etiology not specified; mostly adult sample with 2 children (age range: 14-62y) | Passive listening | Temporal ROI. Count of voxels activated with an extent threshold of $> 10$ voxels. |
| Sabbah et al. (2003) | Individual participant data from article | Left & right: mostly TLE; mixed etiology; adult and pediatric sample (age range: 9-48y) | Semantic fluency | Whole brain ROI. Count of voxels activated above $p < 0.0001$ in ROI |
| Stasenko et al. (2022) | Individual participant data from | Left & right: TLE; mostly HS; adult sample (age range: 22-49y) | Semantic judgement | Frontal + temporal ROI. Count of voxels activated above $p < 0.01$ in ROI |

AGE AT SEIZURE ONSET AND LANGUAGE LATERALIZATION
|  | article |  |  |  |
| --- | --- | --- | --- | --- |
| Sveller et al. (2006) | Correlation coefficient from article | Left TLE; mixed etiology; adult sample (range unknown, mean = 32y) | Verb generation | Frontal + temporal ROI. Count of voxels activated above $p < 0.001$ in ROI |
| Szaflarski et al. (2008) | Individual participant data from article | Left & right; mostly TLE; mostly HS; adult sample with one child (age range: 17-53y). | Verb generation* and semantic decision/tone decision | Frontal + temporal ROI. Count of voxels with z-scores $\geq 2.58$ in ROI |
| Thivard et al. (2005) | Individual participant data from article | Left & right; TLE: mostly HS; adult sample (age range: 18-55y) | Semantic fluency* and story listening | Frontal ROI. Count of voxels activated above $p < 0.05$ (FWE) in ROIs |
| Tivarus et al. (2012) | Individual participant data from article | Left only; mostly TLE; mostly HS; adult sample (age range: 39-69y) | Conjunction of four language tasks: verb generation, semantic decision, definition naming and passive sentence reading | Frontal + temporal ROI. Count of voxels with z-scores $\geq 2.3$ in ROI |
| Trimmel et al. (2019) | Individual participant data from author | Left & right; TLE; etiology not specified; adult sample (age range: 19-58y) | Phonemic fluency* and auditory naming<sup>†</sup> | Frontal ROI. Count of voxels activated above $p < 0.05$ (FWE) in ROIs |
| van der Kallen et al. (1998) | Individual participant data from article | Left; mostly temporal; etiology not specified; adult sample (age range: 26-49y) | Verb generation | Whole brain ROI. Count of voxels activated above $p < 0.0001$ in ROI. |
| Voets et al. (2006) | Individual participant data from article | Left only; TLE; mostly HS; mostly adult sample (age range: 15-53y) | Phonemic fluency | Frontal ROI. Activation change in voxels with maximum activation ROI |
| Wilke et al. (2011) | Individual | Left & right; FLE and TLE; mixed | Expressive letter task* and | Frontal ROI. LI toolbox: Voxel |

AGE AT SEIZURE ONSET AND LANGUAGE LATERALIZATION
|  | participant data from author | etiology; pediatric sample with 2 adults (age range: 5-18y) | receptive beep-stories task | count/value, mean LIs determined across different thresholds using bootstrapping methods in ROIs |
| --- | --- | --- | --- | --- |
| Yuan et al. (2006) | Individual participant data from article | Left & right; mixed location and etiology, many MRI negative; pediatric sample with 2 adults (age range: 8-19y). | Verb generation | Frontal ROI. Count of voxels activated above threshold calculated from the mean value of the t-statistics for all voxels within ROI |
| CNH sample | Additional sample | Left & right; mixed location and etiology including MRI negative; mostly pediatric sample with 17 adults (age range: 5-23y) | Auditory description decision | Frontal ROI. LI-toolbox: Voxel count/value, mean LIs determined across different thresholds using bootstrapping methods in ROIs |
| GOSH sample | Additional sample | Left & right; mixed location and etiology; pediatric sample with 1 adult (age range: 4-19y) | Verb generation | Frontal ROI. LI-toolbox: Voxel count/value, mean LIs determined across different thresholds using bootstrapping methods in ROI |

\* Indicates the language task used if multiple were reported. <sup>†</sup> Indicates the language task used for the temporal LI if different from the task used for the frontal LI.
fMRI = functional magnetic resonance imaging; FDR = false discovery rate; FWE = family wise error; CNH = Children's National Medical Centre; GOSH = Great Ormond Street Hospital; HS = hippocampal sclerosis; LI = laterality index; ROI = region of interest; TLE = temporal lobe epilepsy.

### 2.6 Measures

#### 2.6.1 Laterality Indices

We measured language lateralization using LIs calculated from task-based fMRI. We used continuous LIs as the outcome measure for all analyses. Where studies reported LIs for multiple ROI or language tasks, we chose to use LIs calculated from frontal ROIs during tasks which most robustly activated frontal regions (e.g., verbal fluency). Frontal ROI were the most consistently reported across included studies, and have shown to be reproducible (Harrington et al., 2006) and robustly lateralizing in healthy participants (Bradshaw et al., 2017). The ROI and task used for each study are reported in Table 3. 15 studies reported LI calculated in both frontal and temporal ROI and could be included in the multivariate meta-analysis to examine whether ROI moderated the relationship between age at seizure onset and LI. Of these studies, 13 reported temporal LIs for same task as frontal LI, and for two they only reported temporal LIs for an alternative language task.

#### 2.6.2 Age at seizure onset

Studies reported ‘age at seizure onset’ as the age at which habitual seizures occurred. Only two studies also reported an age at first seizure and four reported age at precipitating injury. This was insufficient for meta-analysis.

#### 2.6.3 Lateralization and localization of epilepsy pathology

We coded the lateralization and localization of epilepsy as reported in the articles or shared by study authors. For the additional UK and US samples, the side of epilepsy was coded based on the structural abnormalities from the clinical MRI reports. For all those who went on to have surgery, the side of epilepsy was consistent with the side of surgery. For scans that were reported as “MRI negative”, the side of epilepsy was based on the seizure focus as reported in the clinical electroencephalography reports. Individuals were excluded if no side of epilepsy was reported or if they had a bilateral epilepsy. For the subgroup analyses we coded whether the structural abnormality involved frontal or temporal lobes only.

### 2.7 Analysis strategy

#### 2.7.1 Calculation of correlation coefficients

For most of the studies we calculated the correlation between age at seizure onset and language lateralization. We calculated Pearson correlation coefficients for the correlation between LI and age at seizure onset for all relevant groups (primary analysis: left hemisphere epilepsy, right hemisphere epilepsy; subgroup analysis: left frontal epilepsy, left temporal epilepsy; multivariate analysis: frontal ROI; temporal ROI).

#### 2.7.2 Meta-analyses

All meta-analyses were conducted using the *dmetar, meta* and *metafor* packages in R (Balduzzi et al., 2019; Harrer et al., 2021; Viechtbauer, 2010). A random-effects model with a Restricted Maximum Likelihood estimator (Viechtbauer, 2005), was used to account for expected heterogeneity in effect sizes (Field, 2001; Hunter & Schmidt, 2000). Between study heterogeneity was explored by examining Cochran’s Q statistic (Cochran, 1950) and the I^2^ statistic (Higgins & Thompson, 2002), with I^2^ values of 25%, 50% and 75% indicating low, moderate and high heterogeneity, respectively. Influence analysis using the leave-one-out method was used to explore the influence of individual studies on the pooled effect sizes and between studies heterogeneity (Viechtbauer & Cheung, 2010). Publication bias was explored by visual examination of contour-enhanced funnel plots and by assessing their asymmetry using Egger’s tests. If publication bias was present, the Duval and Tweedie trim-and-fill method was applied to examine the pooled effect size after adjusting for publication bias.

Studies were identified as outliers if there was no overlap between the 95% confidence intervals of that study and the 95% confidence intervals of the pooled effect size. The meta-analysis was rerun with these outliers removed

#### 2.7.3 Exploratory analyses of individual participant data

Due to the unexpected volume of individual participant data available (n=1157), additional exploratory analyses were conducted which were not reported in the preregistration. We used multilevel models to examine whether language lateralization was best predicted by a linear, logarithmic or categorical coding of age at seizure onset, with study included as a random effect. Due to the negative skew in the residuals of these models we performed an exponential transformation of the LI values to reduce the skew. All data visualization used untransformed LI values. For the logarithmic predictor model, we added a constant of one to the age at seizure onset and then performed a logarithmic transformation. For the categorical model, we coded age at seizure onset as ‘early’ (< 6 years) versus ‘late’ (≥ 6 years), given that 6 years of age has been suggested as a cut-off, after which language networks become more specialized and plasticity declines (Berl, Mayo, et al., 2014; Olulade et al., 2020; Saltzman-Benaiah et al., 2003). We compared the Akaike information criterion (AICs) and Bayes Factor (BF) of the three model and a baseline model with random effects only and ran chi-square tests, to choose the model with best fit. These analyses allowed us to more directly compare linear and sensitive period models of plasticity.

### 2.8 Quality of reporting

The quality of reporting for each study was assessed using a modified version of the ‘Patients’ section of the Evidence-Based Neuropsychology checklist (Hrabok et al., 2013) which can be found in Table 4. Studies were assessed based on four criteria which determined whether studies reported sufficient information on demographic and epilepsy-associated variables as well as inclusion/exclusion criteria and patient selection. Studies were given a ‘yes’ for each criterion if they reported the relevant information for each participant, ‘partial’ if they reported this information on a group-level only, or ‘no’ if they failed to report this information.

**Table 4.** Quality of reporting criteria (adapted from Hrabok et al., 2013)

| Criteria |
| --- |
| 1. Were patients described demographically (i.e., age & sex)? |
| 2. Were the patients described clinically? (i.e., age at seizure onset, epilepsy pathology and location) |
| 3. Were the inclusion/exclusion criteria specified? |
| 4. Was patient selection specified (e.g., consecutive patients included)? |

## 3. Results

### 3.1 Sample characteristics

The total sample included a similar number of adults (55%) and children. Many language tasks were used, but the largest proportion of individuals had laterality indices reported for fluency tasks (including phonemic and semantic fluency, and verb generation; 54%), auditory description tasks (17%), semantic decision tasks (10%) listening tasks (7%) or a conjunction of multiple tasks (6%), with all other tasks being used in less than 5% of the sample each. Most individuals had a laterality index calculated in a frontal ROI (66%), compared to temporal (2%), frontotemporal (27%) or whole brain ROI (5%).

### 3.2 Primary meta-analysis on total sample

There was a correlation between an earlier age at seizure onset and greater atypical language lateralization across the total sample (r=0.1, p=.005, k=58, n=1240). The between-study variance was estimated at tau^2^=0.0026 (95% CI: 0-0.0596) with an I^2^ value of 8.1% (95% CI: 0-33.7%) and a non-significant Q statistic (Q=62.03, p=.302), indicating low heterogeneity in the sample. Influence analysis indicated that effect size was not substantially influenced by individual studies with pooled effect sizes ranging from 0.08 to 0.11, all of which indicated a significant effect. Egger’s regression test for funnel plot asymmetry was non-significant (p=.109) indicating a lack of evidence of publication bias (see the contour-enhanced funnel plot in Supplementary Fig. 1). After the removal of three identified outliers (left hemisphere samples: Appel et al., 2012; Norrelgen et al., 2015; right hemisphere samples: Cano-Lopez et al., 2018), the correlation remained (r=0.12, p<.001, k=55, n=1199).

### 3.3 Subgroup analysis 1: Left vs right hemisphere epilepsy

There was a correlation between age at seizure onset and LI in the left (r=0.1, p=.015, k=37, n=919) but not right hemisphere groups (r=0.07, p=.319, k=21, n=321). There was no difference in the correlation between groups (Q=0.20, p=.656). The pooled effect size and correlation coefficients of the individual samples for the left and right hemisphere groups can be seen in Fig. 2. After the removal of the previously identified outliers, the correlation was significant in both the left (r=0.12, p<.001, k=35, n=887) and right hemisphere groups (r=0.12, p=.014, k=20, n=312) and there remained no difference in the correlation between groups (Q=0.00, p=1).

**Fig. 2.**
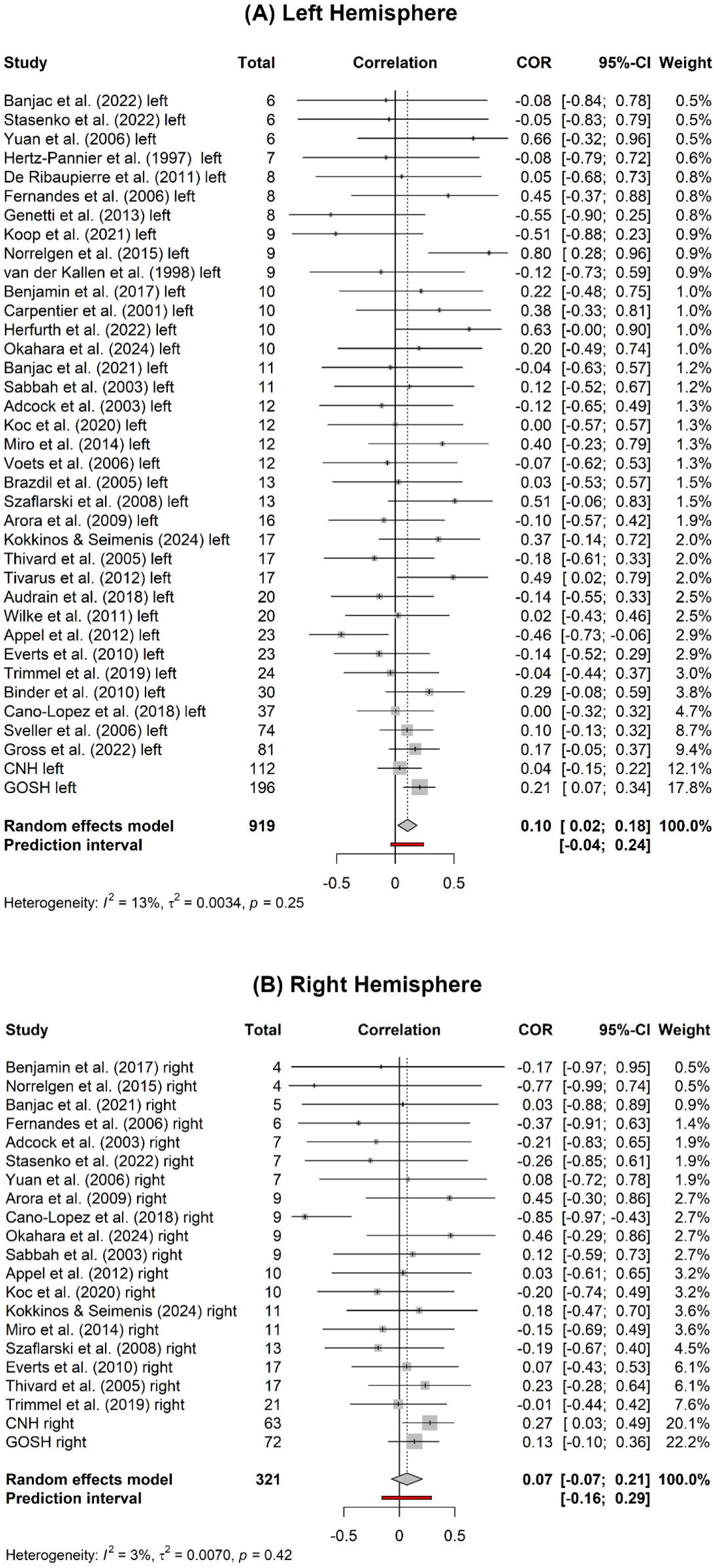
Forest plots showing the individual effect sizes for each study and pooled effect size (dashed line) of the correlation between age at seizure onset and LI in the left (A) and right hemisphere (B) epilepsy groups.

### 3.4 Subgroup analysis 2: Left frontal vs temporal epilepsy

There was no significant correlation between age at onset and LI in the left frontal group (r=0.12, p=.235, k =6, n=82) or left temporal epilepsy groups (r=0.09, p=.123, k=32, n=535), and there was no significant difference between the two groups (Q=0.05, p=.820). After the removal of two outliers (temporal samples: Appel et al., 2012; GOSH), the results remained consistent (temporal: r=0.09, p=.073, k=30, n=447; Q=0.09, p=.766).

### 3.5 Multivariate meta-analysis: comparison of frontal and temporal ROIs for LI calculation

The multivariate meta-analysis revealed no significant moderating effect of the ROI chosen for LI calculation (frontal vs temporal) on the correlation between age at onset and LI (QM=0.63, p=.730). There was no significant correlation between age at onset and LI when using a frontal (r=0.07, p=.478, k=15, n=491) or temporal ROI (r=0.05, p=.482, k=15, n=491).

### 3.6 Comparison of linear, logarithmic and categorical models

Individual participant data were available for 1157 participants. Separate multilevel models were used to compare a linear, logarithmic and categorical coding of age at seizure onset in predicting language lateralization. Given that in our primary analyses, there was no differences in the correlation between age at seizure onset and language lateralization in left and right epilepsy groups, we included left and right hemisphere groups together in this analysis. All three models were significantly better at predicting language lateralization than a model containing only the random effects (AIC: 2306; all p<.01). The individual estimates for the onset predictors in each model can be seen in Table 5. The logarithmic model had the lowest AIC (2287), followed by the categorical (2294) and linear models (2299). There was strong evidence that the logarithmic model is a better fit than the linear model (BF=350) and categorical model (BF=36) and weak evidence that the categorical model is a better fit than the linear model (BF=10). The different models overlayed on the raw data can be seen in Fig. 3.

**Table 5.** Individual coefficients for the fixed effects of each multilevel model.

| Model | Variable | Estimate | t | p value |
| --- | --- | --- | --- | --- |
| **Linear** |  |  |  |  |
|  | ASO | 0.01 | 2.99 | .003** |
| **Logarithmic** |  |  |  |  |
|  | Log(ASO) | 0.11 | 4.57 | <.001*** |
| **Categorical** |  |  |  |  |
|  | Early ASO (< 6 years) | -0.15 | -3.66 | <.001*** |

ASO = age at seizure onset; \* $p < 0.05$ , \*\* $p < .01$ , \*\*\* $p < .001$

**Fig. 3.**
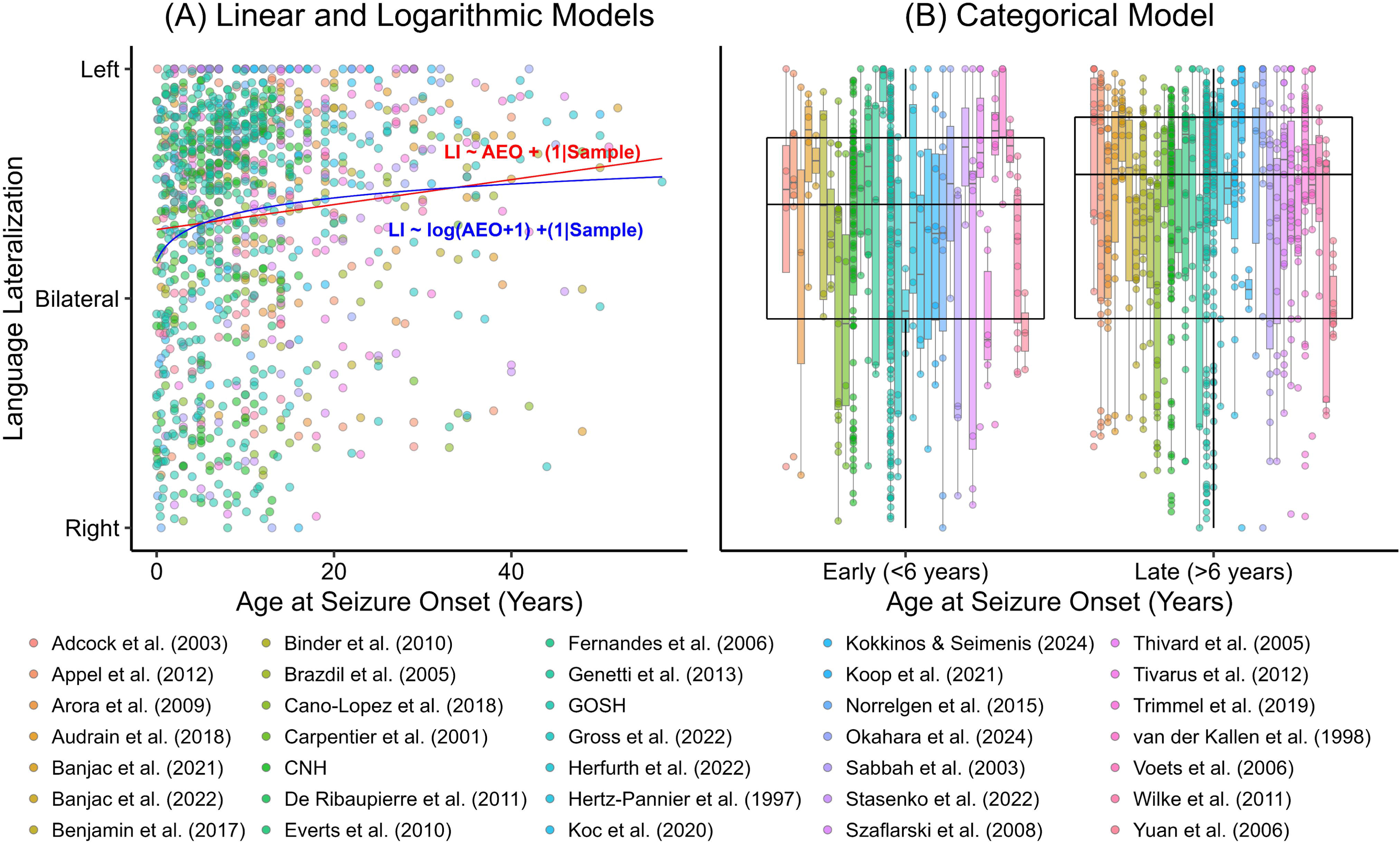
Relationship between age at seizure onset and language lateralization as characterized by linear (red), logarithmic (blue) and categorical multilevel models.

### 3.7 Quality of reporting

Each published study was assessed on four criteria adapted from the patient section of the Evidence-Based Neuropsychology checklist. The proportion of studies reporting relevant information for each criterion is demonstrated in Fig. 4.

**Fig. 4.**
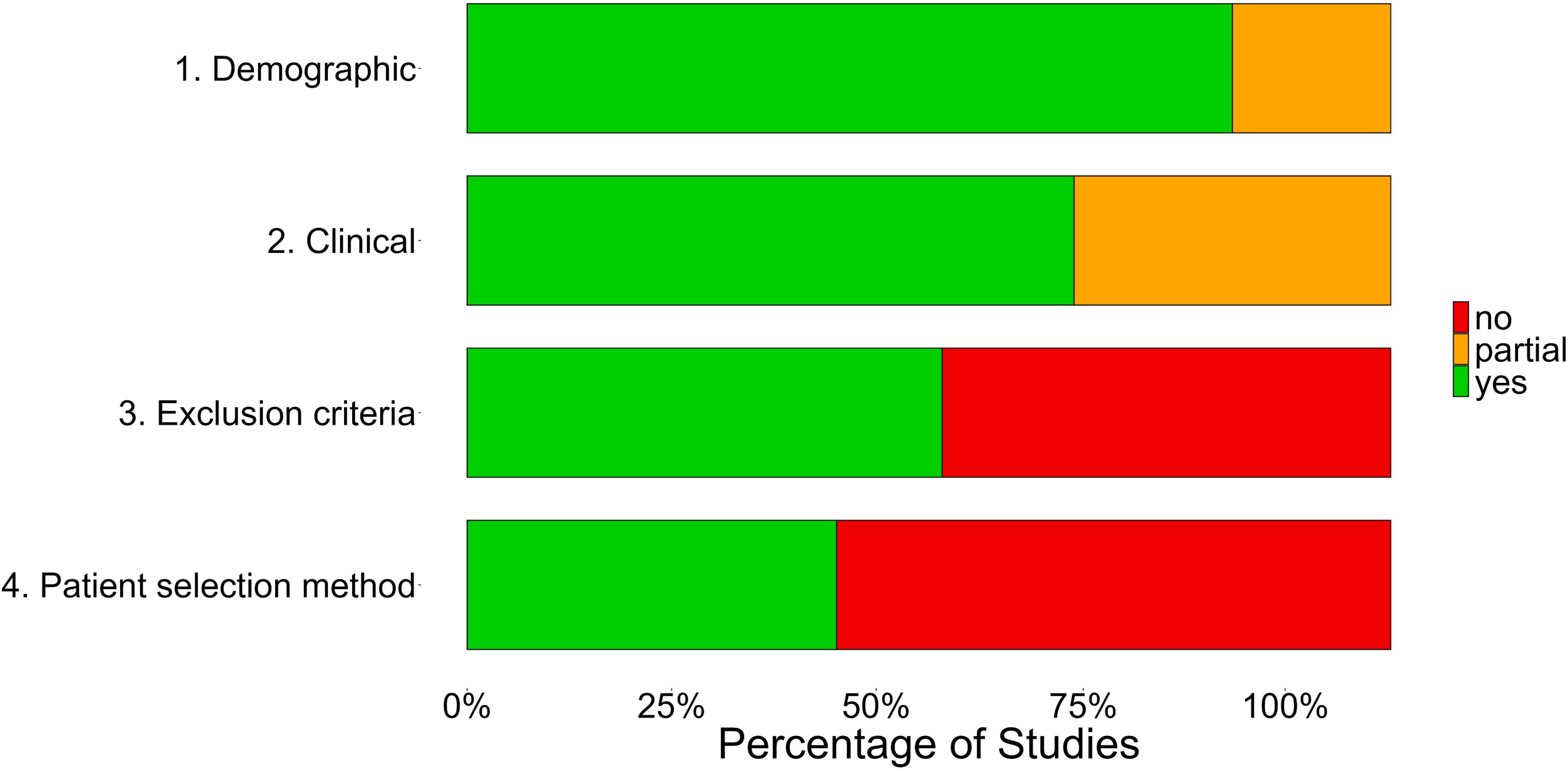
Proportion of included studies reporting relevant patient and study characteristics.

## 4. Discussion

In our meta-analyses we demonstrated a small but significant correlation between an earlier age at seizure onset and greater atypical language lateralization. There was no difference in the correlation between left and right hemisphere epilepsy groups, and the correlation was significant in both groups after the removal of outliers. This result was unexpected in the right hemisphere epilepsy group, given that the reorganization of language functions to the right hemisphere has often been associated with the presence of structural lesions or seizures in the left hemisphere only (Adcock et al., 2003; Berl et al., 2005; Carpentier et al., 2001; Liégeois et al., 2004). It is possible that an early onset of epilepsy in the left or right hemisphere may similarly disrupt typical trajectories of language lateralization, leading to atypical representation regardless of the side of epilepsy. This appears consistent with the contribution of the RH in early language development which has been demonstrated to decline over childhood, while LH involvement remains consistent (Olulade et al., 2020). Although early right hemisphere injury has been shown to cause delays in language development (Thal et al., 1991; Trauner et al., 2013), it is still unclear what effect, if any, such an injury has on the development of lateralized language networks.

### 4.1 Evidence for a nonlinear relationship

We found strong evidence that a logarithmic model fitted the data better than a linear model, indicating that there may be a greater likelihood of reorganisation at an earlier age, which rapidly reduces over early childhood before stabilising. Presumable this would reflect an exponential decline in plasticity over the first several years of life, compared to a more gradual linear decline in later childhood. This provides some support for the sensitive period hypothesis. However, there was strong evidence that the logarithmic model was a better fit than the categorical model, suggesting that there is not a definitive cut-off point for language reorganization, and that after six years of age there may still be a subtle influence of epileptic pathology/activity on language lateralization.

### 4.2 Subtle effect as opposed to robust clinical indicator

Regardless of how it was characterised, the relationship between age at seizure onset and language lateralization was subtle and diminished in sub-analyses with smaller sample sizes. This indicates that an earlier age at seizure onset may not be a good clinical marker of language lateralization at the individual patient level. There may be other factors which have a greater influence on reorganization, such as the presence of early acquired injuries (Duchowny et al., 1996; Gaillard et al., 2007; Rathore et al., 2009). While all studies included in our analyses reported the age at onset of habitual seizures, fewer reported the age at first seizure or precipitating injury. In patients where the onset of habitual seizures is preceded by an earlier developmental event such as an acquired brain injury, the age at habitual seizure onset may not accurately reflect the point of initial reorganization of language functions.

Consequently, there could be a stronger relationship between language lateralization and the age at precipitating event or first seizure. However, previous research suggests that both age at habitual seizures and precipitating events are associated with language lateralization (Rathore et al., 2009; Woermann et al., 2003), and in fact the former association may be stronger (Springer et al., 1999).

### 4.3 Limitations

Studies included in this meta-analysis used different language tasks and ROIs for the calculation of language lateralization, and this may explain some of the heterogeneity in effect sizes between studies. Where possible, we extracted LI for frontal ROI, however, a third of the sample included LIs from temporal, fronto-temporal, or whole brain ROI. Our multivariate meta-analysis indicated that ROI choice did not have a significant moderating effect on the correlation between age at seizure onset and LI. This is perhaps unsurprising, given that most of the temporal LIs were calculated for the same tasks as frontal LIs were calculated for. It may be that temporal LIs calculated for different, and more traditionally receptive, tasks would differ from frontal LIs calculated using expressive tasks.

Our findings may have limited generalisability to other epilepsy samples. Studies included in our meta-analysis may be biased in their epilepsy sample, as many patients will have language fMRI as part of a presurgical evaluation for resective or disconnective surgery. It is likely that this surgical cohort would vary compared to a non-surgical focal epilepsy cohort, most obviously in terms of seizure burden, as these patients tend to have medication-resistant seizures. It is therefore possible that these findings do not generalise to samples with focal epilepsy which is well controlled by medication. As can be seen in Fig. 4, more than half of the included studies failed to report their exclusion criteria or patient selection procedures, which makes it difficult to determine what other biases might be present in these groups.

### 4.4 Conclusions

In our meta-analysis we identified a small but significant correlation between age at seizure onset and language lateralization in a large focal epilepsy sample of 1254 individuals, regardless of the side of epilepsy pathology. This relationship was best characterized with a logarithmic curve, likely reflecting an exponential decline in plasticity over early childhood, but with no clear definitive cut-off for reorganization. Given that the effect of age at seizure onset on language lateralization was subtle and only present in large samples, an early age at seizure onset would not serve as a good indicator of atypical language lateralization on the individual patient level.

## Funding

FP is supported by the Child Health Research Studentship, funded by the Child Health Research Charitable Incorporated Organisation and spent 8 weeks at the Children’s National Hospital in Washington, DC, funded by The Charlotte and Yule Bogue Research Fellowships in Honour of Sir Charles Lovatt Evans and A.J. Clark. MHE is supported by the Child Health Research Studentship, funded by the National Institute for Health and Care Research Great Ormond Street Hospital Biomedical Research Centre (NIHR GOSH BRC) and the Sigrid Jusélius Foundation. LNS is supported by a K23 grant (K23 NINDS NS093152).

## Data Availability

The summary statistics and meta-analyses script can be found on the Open Science Framework. Data sharing requests should be directed to the corresponding author of the original article.

https://tinyurl.com/bdctb3t2

## Acknowledgements

This work was supported by the National Institute for Health and Care Research Great Ormond Street Hospital Biomedical Research Centre (NIHR GOSH BRC). We are ever so grateful to the authors who responded to our request for data, whether those data were available or not. Thank you for supporting our work.

## Footnotes

1 The original search was conducted on May 19^th^ 2022 and was topped up on August 14^th^ 2024, when additional MeSH and Embase terms were added.

## Notes

### Competing Interest Statement

The authors have declared no competing interest.

### Author Declarations

Ethical approval was granted by the Great Ormond Street Hospital clinical audit department as a service evaluation (No. 1443, extended), according to the guidelines set by NHS Research Ethics Committee Review. Participants from Children's National Hospital (CNH) were part of clinical or research protocols with different aims related to pediatric epilepsy and language development, but all were approved by the Institutional Review Board at CNH. Parents provided informed written consent and all children gave assent to participate.

