## Supplementary Material for "Is an earlier onset of focal epilepsy associated with atypical language lateralization? A systematic review, meta-analysis and new data"

### Supplementary Materials

#### 1.1 Exclusion of individual participants

*Supplementary Table 1. List of studies with individuals removed for the meta-analysis*

| Study | Participants excluded, reason |
|---|---|
| Arora et al. (2009) | Missing LI for relevant task (9), no specific age at seizure onset (3), bilateral epilepsy (1), epilepsy not lateralized (1), prior resection (1) |
| Banjac et al. (2022) | Overlapping with Banjac et al. (2021) (12) |
| Benjamin et al. (2017) | Postoperative fMRI only (4), poor quality fMRI (3) |
| Hertz-Pannier et al. (1997) | Epilepsy not lateralized (1) |
| Herfurth et al. (2022) | Missing LI for relevant task (10) |
| Kokkinos & Seimenis (2024) | Epilepsy not lateralized (2) |
| Norrelgen et al. (2015) | Epilepsy not lateralized (4), insufficient activation for LI calculation (2) |
| Okahara et al. (2024) | Epilepsy not lateralized (2) |
| Szaflarski et al. (2008) | Postoperative fMRI only (2) |
| Thivard et al. (2005) | Insufficient activation for LI calculation (2) |
| Trimmel et al. (2019) | Missing LI for relevant task (1) |
| van der Kallen et al. (1998) | No specific age at seizure onset (3) |
| Yuan et al. (2006) | Epilepsy not lateralized (5) |
| Wilke et al. (2011) | Missing LI for relevant task (4) |

#### 1.2 Meta-analysis with onset < 18 years of age

There was no significant correlation between age at seizure onset and language lateralization in the sample with onset before 18 years of age (r=0.1, p=.05, k=51, n=897). Influence analysis indicated that effect size was substantially influenced by individual studies with pooled effect sizes ranging from 0.07 to 0.11. Egger’s regression test for funnel plot asymmetry was significant (p=.049). After applying the Duval and Tweedie trim-and-fill method to adjust for publication bias, the correlation became significant (r=0.14, p=.005). Four samples were identified as outliers due to the lack of overlap between the 95% confidence intervals of these studies and the pooled effect size (left samples: Hertz-Pannier et al., 1997; Norrelgen et al., 2015; Szaflarski et al., 2008; right sample: Koc et al., 2020). After the removal of these outliers, the correlation became significant (r=0.1, p=.009, k=47, n=869).

There was no significant correlation between age at seizure onset and LI in the left (r=0.08, p=.246, k=33, n=643) or right hemisphere group separately (r=0.13, p=.200, k=18, n=254), and no difference in the correlation between groups (Q=0.30, p=.585). After the removal of the four previously identified outliers, the correlation became significant in the right (r=0.15, p=.031, k=17, n=248) but not the left hemisphere group (r=0.07, p=.175, k=30, n=621). There was still no difference in the correlation between groups (Q=1.12, p=.290).

##### 1.3 Supplementary figures

*Supplementary Fig. 1. Contour-enhanced funnel plot for the total sample meta-analysis.*

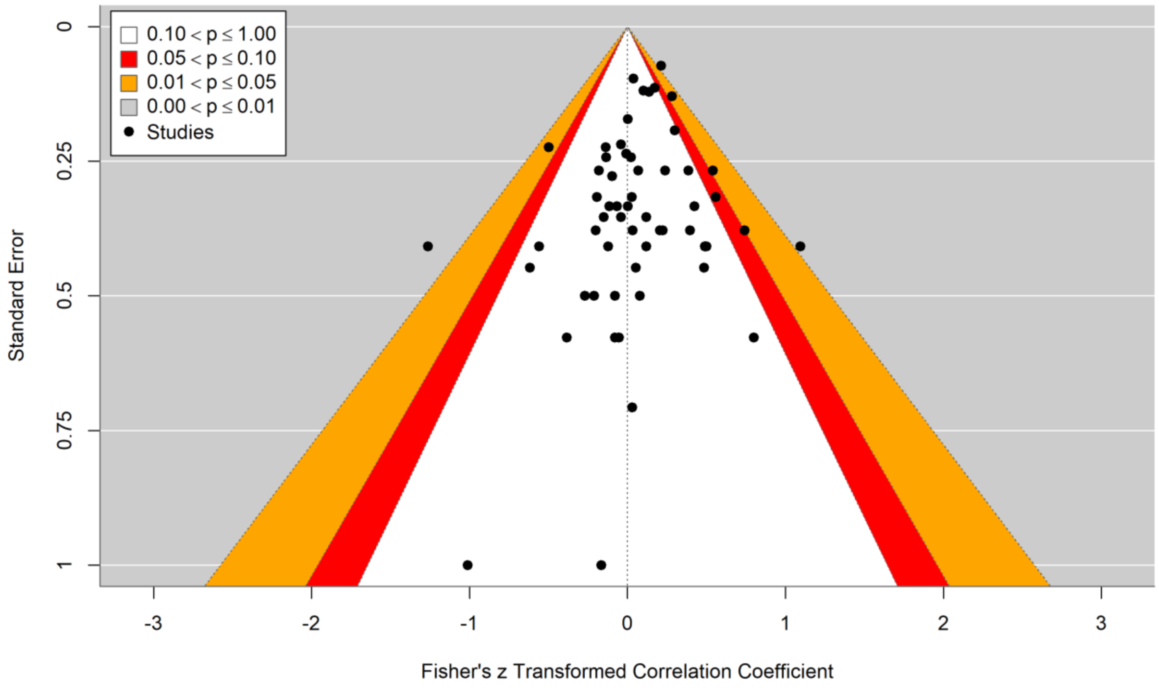
